# Evaluating the Evidence Quality Supporting the Role of Acupuncture for Herpes Zoster Pain: An Umbrella Review of Systematic Reviews and Meta-analyses

**DOI:** 10.1101/2022.09.30.22275406

**Authors:** Jianhong Li, Kun Liu, Wenjia Ruan, Guohua Lin

## Abstract

**Objective:** Acupuncture is one of the most effective means to treat herpes zoster, especially during the acute stage. It can shorten the duration of pain, with a low incidence of residual neuralgia. The aim of this review was to assess the methodological and evidence quality of systematic reviews and meta-analyses on acupuncture for herpes zoster pain and provide clinical suggestions.

**Methods:** Eight databases were searched from their inception to Feb 2022. Methodological quality was evaluated by using the Assessment of Multiple Systematic Reviews 2, and the Grading of Recommendations Assessment Development and Evaluation was applied to evaluate the quality of the evidence.

**Results:** Nineteen SRs/MAs were included according to the inclusion criteria. Assessment of Multiple Systematic Reviews 2 results showed that the methodological quality was low for 16 studies and very low for 3 studies. According to the Grading of Recommendations Assessment Development and Evaluation, 43 outcome indicators were evaluated in this review, and the results showed that the evidence quality was high in three, medium in twelve, low in twenty-three, and very low in five. Compared with drugs for herpes zoster, acupuncture can reduce pain and the incidence of postherpetic neuralgia. Acupuncture at Jia ji acupoint was generally one of the most effective methods.

**Conclusions:** Our study showed that fire needle, acupuncture at Jia ji, pricking collaterals and bloodletting are beneficial therapies for the clinical practice of herpes zoster. However, the methodological and evidence quality of the included studies were low. More rigorous design and comprehensive investigations by systematic reviews and meta-analyses are needed.

**Strengths and limitations of this study:** This study is a comprehensive review to assess the methodological and evidence quality of recent SRs/MAs on acupuncture for herpes zoster pain, we mainly focused on pain symptoms, for example VAS score, time for pain relief, incidence of PHN, with qualitative analysis of efficiency. Only systematic reviews published in Chinese and English were included, so the conclusions of this study may not be generalizable. Some gray documents may be omitted. Subjective factors could not be completely eliminated during screening and evaluation.

PROSPERO registration number CRD42022304247.

## Introduction

Herpes zoster (HZ) is caused by varicella-zoster virus infection, which is a DNA virus with neurophilic characteristics (1,2). After the first infection, it is hidden in the sensory ganglion. When immunity decreases due to infectious diseases and long-term immunosuppressive treatment, the latent virus can reactivate, causing inflammation and necrosis of the affected ganglion. HZ can occur at any age but is most common in elderly people (3-5). The natural course is 2-3 weeks. It is characterized by a banded distribution of blisters along one side of the skin nerve, often accompanied by severe and lasting neuralgia, local lymph node pain and progressive aggravation. The average incidence of acute HZ is 3-5 million people per year. Acupuncture can relieve acute pain, prevent or alleviate acute or chronic complications such as post-herpetic neuralgia (PHN). Drug therapy is the first-line treatment at present, including antiviral drugs (recommended for HZ patients over the age of 50), neurotrophic drugs and painkillers such as lidocaine patches and topical capsaicin (6). Conventional drugs such as antiviral drugs can shorten the healing time, but long-term excessive use may lead to side effects such as hemolytic anemia, thrombocytopenia or renal insufficiency. In addition, there are many taboos related to the treatment of elderly and frail patients (7).

Acupuncture has been used to treat HZ in China for a long time, and acupuncture is an effective treatment in reduction in pain, especially HZ-related pain. Chinese medicine experts generally recommend acupuncture or acupuncture-based methods such as fire needle, electroacupuncture, local acupuncture, pricking and bloodletting, cupping, moxibustion and other treatments (1) during the acute stage of HZ (8). Acupuncture treatment of HZ is listed as a grade I in the “Modern Acupuncture and moxibustion Disease Spectrum” (9). Systematic reviews and meta-analyses (SRs/MAs) on acupuncture treatment for HZ have appeared since 2007. However, these SRs/MAs have not been systematically evaluated, we conduct this umbrella review to assess these studies to provide recommendations for conducting high-quality SRs/MAs.

## Methods

### Data retrieval strategy

The following databases were searched from their inception to Feb 2022: Embase, PubMed, Embase, Cochrane Library, Chinese Biomedical Literature Database (CBM), Chinese Medical Current Content (CMCC), China Science and Technology Journal Database (VIP database), Wan-Fang Database, China National Knowledge Infrastructure (CNKI) and Citation Information by National Institute of Informatics (CiNii). The following keywords were used: acupuncture, acupoint, electroacupuncture, fire needle, spinous bloodletting, auricularpoint, herpes zoster, systematic review, meta-analysis, etc. Detailed retrieval strategies are shown in Table 1.

**Table 1.** Retrieval Strategy.

### Inclusion and exclusion criteria

#### Types of studies

SRs/MAs based on randomized controlled trials (RCTs) and controlled clinical trials (CCTs).

#### Types of subjects

Patients with herpes zoster were enrolled in accordance with the consensus of Chinese experts (3), regardless of sex, age, race, and region.

#### Types of interventions

The treatment group included patients receiving acupuncture, fire needle, puncture and cupping, skin acupuncture or other treatments in combination with the aforementioned treatments; the control group included patients receiving drug therapy (antiviral acyclovir or traditional Chinese medicine, etc.).

#### Types of outcome measurements

The outcome measurements were efficacy (refer to “Guiding Principles for Clinical Study of New Chinese Medicines”), pain evaluation (pain relief time, visual analogue scale VAS), and incidence of PHN. Studies were excluded according to the following criteria: □ repeatedly published literature or review comments; ②non-SRs/MAs; ③clinical trials only for the study of postherpetic neuralgia; ④conference papers, abstracts, dissertations, etc.; ⑤documents with incomplete content and unable to extract data.

### Data extraction

Two trained researchers independently screened the literature, extracted and cross-checked the data. In cases of disagreement, the third reviewer was consulted to help judge and make up for the lack of data as much as possible. The data extraction table was generated in Excel, including the title, author, year of publication, number of studies, number of cases, interventions, outcome measurements. The consistency among evaluators was evaluated by the Kappa value.

#### Evaluation of the methodological quality of the included studies

Two evaluators used AMSTAR-2 (10) to evaluate the methodological quality of the included studies under the premise of hiding the author. The AMSTAR-2 contains 16 items in total, of which items 2, 4, 7, 9, 11, 13 and 15 were key items to evaluate the included studies. Each was evaluated as “yes”, “no” or “partial yes” according to the evaluation criteria. According to the evaluated result, the quality was defined as high, medium, low, or very low.

#### Evaluation of the quality of evidence of the included studies

Two evaluators focused on efficiency, VAS score and incidence of PHN and applied GRADE (11) to evaluate the evidence quality from the following five aspects: research limitation, inconsistency, indirectness, imprecision and publication bias. VAS scores were recombined and analyzed (12).

#### Evaluation of the curative effect

The quantitative analysis results included in the systematic evaluation outcome measurements are summarized in the form of relative risk (RR), odds ratio (OR), standardized mean difference (SMD), mean difference (MD), 95% confidence interval (CI) and efficiency.

## Results

### Results of the literature search and selection process

According to the retrieval strategy, a total of 77 related articles were selected, 36 duplicates were removed, 3 studies were excluded after record screening, and 19 studies were included (13-31). The entire process and results are shown in Figure 1.

**Figure 1.** Flow chart of the literature selection process.

### Description of the included reviews

All included studies were published from 2007 to 2022, including 5 English articles, and the rest were in Chinese. The 318 included studies involved 13490 cases. Among them, 3 articles (14-16) included RCTs and CCTs, and 16 articles (13,17-31) were RCTs. Thirteen articles (13,15,18,19,21-23,25-31) used the bias risk assessment tool recommended by Cochrane. Sixteen articles (13-22, 24-26) described two evaluators for data extraction and analysis. The basic characteristics of the included studies are shown in Table 2.

**Table 2.** Basic characteristics of the included studies.

### Evaluation results of the AMSTAR-2 and GRADE studies

#### Methodological quality of the included SRs/MAs

In the included SRs/MAs, 3 (13,24,25) were rated as medium quality, and 16 (14-23,26-31) were rated as low quality. Only item 1 was evaluated in 19 studies, and item 10 was evaluated in 19 studies; item 16 was unevaluated in 18 studies. The results are shown in Table 3.

**Table 3.** Results of AMSTAR-2 evaluation.

#### Quality of the evidence from the included SRs/MAs

In the included SRs/MAs, 43 outcome indicators were evaluated in this review, and quality of the evidence was high in three, medium in eleven, low in twenty-three and very low in five. Research limitations was the most commonly degraded item, followed by imprecision, inconsistency, publication bias and indirectness. Detailed information is shown in Table 4.

**Table 4.** Results of the GRADE evaluation.

#### Main outcome indicators and levels of evidence

##### Efficacy of acupuncture for the treatment of herpes zoster

Referring to “Guiding Principles for Clinical Study of New Chinese Medicines”, efficacy rate was defined as partial regression of skin lesions, improvement in clinical symptoms, and 70% > syndrome score reduction ≥50%. Twelve systematic reviews (13-14, 16, 19, 21, 23, 26) evaluated the efficacy rate. The curative effect of the experimental group was better than the control group, and the results are shown in Table 3.

The rate of acupuncture efficacy for attenuating pathological changes and improving pain was more than 30% compared with conventional drugs (13), and the curative effect of the experimental group was better than that of the control group (14). There was a significant difference between the Jia ji acupoint and control groups, RR=1.07, 95% CI (0.98∼1.16) (16).

The effect of the treatment group was better than that of the control group, and the difference was significant (21). The overall effect of fire needles on the treatment of herpes zoster was better than medicine (23). In the study of Geng (27), all 17 studies reported that the efficacy of the blood puncture and cupping group was better than that medicine group, and the difference was statistically significant.

Regarding the definition of efficacy, Coyle ME (13), Fu (15), Wang (20), Lin (21), Shi (22), and Li Dong (23) did not specifically describe the efficacy rate.

#### Secondary outcome indicators and evidence levels

##### Pain relief time

Six systematic reviews (13,14,18,19,22,25) evaluated the time of relieving pain. There were significant differences in pain relief between acupuncture and valaciclovir combined with vitamin B1 and saline application; the pain in the acupuncture group disappeared 6.59 days earlier than that in the oral and local use of acyclovir group (13); the electroacupuncture group could relieve pain 13 days earlier (14); the analgesic effect of the treatment group was better than the control group (18); and fire needle could relieve pain 3 days earlier (19). Pain relief time in the fire needle group was significantly shortened (22). In the experimental group, pain relief time was significantly shorter than that in the control group (25).

##### VAS score

Eight systematic reviews (14-16, 17, 22, 24-26) evaluated VAS scores. The VAS score in the experimental group was lower than that in the control group (14, 25, 26); the electroacupuncture score was mostly lower than that medicine group (15), and the difference in VAS score between Jiaji acupoint treatment and carbamazepine, acyclovir and routine acupuncture control groups was statistically significant (16). The differences in VAS score in the acupuncture group was higher than that in medicine group (17). Compared with medicine, fire needle was not exact in relieving the pain of herpes zoster, I^2^=93% (23). VAS scores after treatment mainly by puncture and cupping were lower than those in the antiviral group (31). Lower VAS scores were observed in the fire needle group (27). Acupuncture combined with puncture and cupping is more advantageous in reducing VAS score (33). Acupuncture can alleviate pain significantly (24). The fire needle group was superior to medicine group (30). There was no significant difference between the treatment group and the control group (29). In this study, 30 clinical studies involved in 8 systematic reviews of VAS scores (32-60) were reanalyzed by Revam5.3 software. The results are shown in Figure 2.

**Figure 2.** Forest map of the VAS score after acupuncture-based treatment for herpes zoster.

##### Incidence of PHN

Fourteen systematic reviews (13,14,19,20-27,29-31) evaluated the incidence of PHN. The results are shown in Table 4. One month after the disappearance of the rash, there was a statistically significant difference in the incidence of PHN, but there was no significant difference, 3 months after the disappearance of the rash, RR=0.29, 95%CI (0.16, 0.53), I^2^=0% (13). The differences in the incidence of PHN was significant 30 days after treatment, RR=0.23, 95%CI (0.11,0.51), P=0.0002, and 60 days after treatment, RR=0.33, 95%CI (0.12, 0.91), RR=0.33, 95%CI, 0.03 (22). In addition, at 30 days after treatment, there was a significant difference in the incidence, RR=0.23, which was lower in the fire needle group than in medicine group (19).

#### Safety of acupuncture treatment for herpes zoster

Six systematic reviews (13,17,19,20,28,29) evaluated adverse events, and Coyle ME (13) reported subcutaneous hematoma and hemorrhage in the experimental group. Zhao (17) did not mention whether there were side effects. Wang (20) discussed the adverse reactions of fire needles, including dizziness and fatigue and a mild decrease in white blood cells. Wang (19) also reported 1 case of skin erythema with pruritus, 3 cases of gastrointestinal reaction and 3 cases of diarrhea in the group of fiery needle combined with red light. Only one of the 23 RCTs included in Yin (28) reported adverse reactions to electroacupuncture at the Jiaji acupoint.

## Discussion

### Summary of the evidence

The clinical effect of acupuncture for HZ pain is certain. The AMSTAR-2 results showed that the methodological quality was low for 16 studies and very low for 3 studies. According to GRADE, 43 outcome indicators were evaluated in this review, and the evidence quality was high in three, medium in twelve, low in twenty-three, and very low in five. The methodological and evidence quality of Coyle ME is the highest (13). Three papers included both RCT and CCT studies, which affected the strength of evidence (15-17).

Defects in study design and implementation, such as a lack of registration, relaxed research criteria, and potential risk of bias, such as repeated publication and inclusion, may affect the authenticity and reliability of the results (12). The downgrading of inaccuracy for GRADE, for example the hidden research distribution scheme, lack of blind method, withdrawal consent situation, less than 400 total cases, wide confidence interval, invalid values, and small sample size (16,23). Mixed intervention methods, publication bias, and less overlap of confidence intervals, the value of I^2^ was large may impact on clinical significance (18). A preliminary plan, register on the PROSPERO platform or Cochrane website, funding information report, describe the method of data extraction and the process of obtaining or confirming the data from the author are recommended. Conduct the application of AMSTAR-2, GRADE and other evaluation tools (12) to objectively improve the reporting and quality of SAs/MAs (66).

### Implications for practice and research

Acupuncture has broad prospects in the treatment of pain-related diseases, including HZ. For the form of acupuncture, acupuncture methods at Jia ji point may be superior and safe compared with Western medicine (15,28). Pricking collaterals, bloodletting and fire needles (13-14, 16, 19, 21, 23, 24, 25) can shorten the time to pain relief and reduce the incidence of PHN earlier (27-29). The A shi (tender point around the rashes), EX-B2 (Jia ji), and SJ6 (Zhi gou) acupoints are commonly adopted in dermatology clinic. Electroacupuncture at the Jiaji acupoint can shorten the herpes healing time and relieve pain (64). Acupuncture combined with cupping bloodletting puncture is superior to conventional medicine in improving the total efficacy rate, reducing VAS score, shortening the scab time and reducing the incidence of PHN (21,30,31,33). Fire needle therapy is suggested as one of the routine treatments for herpes zoster (21,23,27).

### Limitations

Only systematic reviews published in Chinese and English were included, so the conclusions of this study may not be generalizable. Some gray documents may be omitted. Subjective factors could not be completely eliminated during screening and evaluation.

## Conclusion

This study showed that acupuncture could be a beneficial therapy for herpes zoster pain, but there is limited credibility due to the low methodological and evidence quality of the included studies. More rigorous design and comprehensive investigations by SRs/MAs are needed to provide strong evidence for convincing conclusions.

## Data Availability

All data produced in the present work are contained in the manuscript

